# Representation of Racial and Ethnic Minority Populations in Dementia Prevention Trials: A Systematic Review

**DOI:** 10.1101/2021.05.18.21257411

**Authors:** Ashley R. Shaw, Jaime Perales-Puchalt, Ellie Johnson, Paul Espinoza-Kissell, Mariola Acosta-Rullan, Sarah Frederick, Anneka Lewis, Huan Chang, Jonathan Mahnken, Eric D. Vidoni

**Affiliations:** University of Kansas Alzheimer’s Disease Center, University of Kansas Medical Center, Kansas City, United States; Department of Biostatistics, University of Kansas, Medical Center, Kansas City, United States

**Author notes:** **Corresponding author** Eric Vidoni, PhD; Address: 4350 Shawnee Mission Parkway, Fairway, KS, 66205.

**Keywords:** Alzheimer’s disease, dementia, underrepresented minorities, aging, prevention

## Abstract

Despite older racial and ethnic minorities (REMs) being more likely to develop dementia they are underrepresented in clinical trials focused on neurological disorders. Inclusion of REMs in dementia prevention studies is vital to reducing the impact of disparities in dementia risk. We conducted a systematic review to characterize the number of REM enrolled in brain health and prevention randomized controlled trials (RCTs). RTCs published from January 1, 2004 to April 21, 2020 were included. Participants were normal cognitive adults aged 45 years and older who participated in a Phase II or Phase III U.S. based preventative trial. Analyses were performed to examine differences in trial characteristics between RCTs that did and those that did not report race/ethnicity and to calculate the pooled proportion of each racial/ethnic group in randomized brain healthy prevention trials. A total of 42 studies consisting of 100,748 participants were included in the final analyses. A total of 26 (62%) reported some racial/ethnic identity data. The pooled proportion of REM participants was 0.256 (95% CI, 0.191, 0.326). There is a lack of racial/ethnic reporting of participants and REMs remain underrepresented in brain health prevention RCTs.

## INTRODUCTION

In 2021 it is estimated that 6.2 million older Americans are living with dementia.(1) The population aged 65 years and older is projected to increase from 56 million to 94.7 million by 2060, with much of the growth being affiliated with the aging of Baby boomers.(2) Additionally, the proportion of racial and ethnic minority (REM) older adults is expected to increase among African Americans (9% to 13%) (3) and Hispanics (8% to 21%) (4) by 2060. As the older population continues to rapidly grow it is projected that the number people living with dementia will also increase. Among people 65 years and older African Americans and Hispanics are disproportionally impacted. Previous research has indicated that older African Americans are twice as likely and older Hispanics are 1.5 times as likely to develop dementia compared to non-Hispanic Whites.(1) Despite the growing number of racial/ethnic minorities in the United States and the current disproportionate impact of dementia among these populations, there is a poor representation of these groups in randomized control trials (RCTs) focused on neurological disorders.(5-7) Furthermore, it is estimated that in dementia RCTs, minority participation rates are lower than 5%.(8)

There are several possible reasons why most racial and ethnic minorities are underrepresented in RCTs, including historical unethical practices and cultural barriers. Unethical practices such as the Tuskegee syphilis study in which researchers infected and withheld treatment for syphilis to Black men(9) and more recently the Havasupai Tribe case, in which DNA samples initially collected for genetic markers of type 2 diabetes had been used in several unrelated studies such as schizophrenia, and migration without consent from tribal members(10) have led to mistrust in research field among REMs. Cultural barriers in RCTs have been documented as lack of tailoring to diverse communities, implicit bias, and investigators; limiting participation of REM within studies.(11) Yet, previous research has indicated that REMs are willing to participate in clinical trials when presented with the opportunity and when trial objectives can be translated in a culturally relevant manner,(12) which demonstrates that REMs are not necessarily hard to reach but are rarely reached. Race is a socially constructed category and a proxy for unique psychosocial factors strongly related to dementia (13) that needs to be considered when designing dementia prevention interventions. Because dementia prevention trials have primarily focused on non-Hispanic Whites, progress in research related to characteristics of dementia among REM has been limited. Dementia prevention is likely to be a critical aspect in reducing racial and ethnic disparities.(14) However, disease prevention is not a one-size fits all model and it is imperative that preventative approaches aimed at mitigating risk factors of dementia among REM incorporate culture.

Inclusion of REMs in dementia prevention studies is vital to reducing the impact of disparities and critical for addressing imperative gaps in knowledge. Therefore, we conducted a systematic review to characterize the number of REMs enrolled in brain health and prevention trials.

## METHODS

We searched Ovid MEDLINE, Embase, CINAHL Complete, and clinical trials.gov published in English from January 1, 2004 to April 21, 2020. Minimum race and ethnicity reporting standards were adopted by the National Institutes of Health in January 2002 (NOT-OD-01-053). Our search window allows for the completion and reporting of smaller clinical trial projects initiated following this directive. Additionally, we screened references from eligible studies to determine additional eligible articles to include in the review. A detailed search strategy of this review is available in the supplementary materials.

We included published RCTs that met the following eligibility criteria 1) enrollees with normal cognition ages 45 years and older, 2) Phase II or Phase III randomized controlled trials, 3) at least one explicitly identified cognitive outcome measure, and 4) United States-based trials. We excluded investigational medication trials seeking FDA approval, trials aimed at treatment of existing cognitive impairment, psychiatric-related cognitive trials (i.e. major depression as primary diagnosis), protocol related articles, clinical trials that did not provide results, RCTs with no cognitive outcome examined, retrospective, and secondary articles.

Eight reviewers (ARS, EDV, JPP, EDJ, SIF, PEK, MDA, AL) independently screened all titles and abstracts. Two reviewers (ARS and EDV) cross checked all titles, abstracts, and completed full text-review of all eligible studies following screening. Information was abstracted from all eligible studies: publication information (first author, title, journal, PubMed ID [PMID], year of publication); funding source; demographics of enrollees (total number of participants, average age, number of females, number and type of race or ethnicity as defined by the study); study design data (type of intervention, intervention components, primary language of intervention delivery, cognitive tests used); Percentages of ethno-racial groups in the eligible studies were only included in this review if it was specifically mentioned in the manuscript.

We examined the differences in trial characteristics between RCTs that did and those that did not report race/ethnicity of trial participants using Student’s t-test for continuous variables and X^2^ tests for categorical variables. Study characteristics examined included average proportion female, average age, sample size (mean), type of intervention (i.e. diet/supplement vs exercise vs. cognitive training vs. multi-domain), funding source, non-English language delivery (yes vs. no).

We conducted a meta-regression analysis to calculate the pooled proportion of each racial/ethnic group in randomized brain healthy prevention trials. Our primary outcome was non-White/non-Hispanic which included a composite of the following racial/ethnic groups African American, Hispanic or Latinx (hereafter referred to as Hispanic), Asian, American Indian or Alaskan Native, Native Hawaiian or Pacific Islander, and Other Race, following standard NIH reporting guidance. We conducted a subgroup analysis to assess the heterogeneity across different study factors including type of intervention (diet/supplement, exercise, cognitive training, multi-domain), cognitive tests in intervention (MMSE, Rey Auditory Verbal, other) and funding (public, private, or mixed). The pooled estimate of proportion and 95% confidence (15) interval were calculated using random effects meta-analyses with inverse variance weighting. The Freeman-Tukey double arsine transformation of the proportion was used in the estimation.(16) Analyses were performed using SAS 9.4 and R 4.0.2. We used the metaprop function from the “meta” package in R to calculate the pooled estimate and confidence intervals.(17)

## RESULTS

A total of 4,600 articles were screened: 4385 non-duplicate abstracts identified via our search strategy, and 215 articles manually added. After review of titles and abstracts, 49 underwent a full text review. Of the 49 articles reviewed; 7 were excluded for not being RCTs or being derivative of the primary report (n=6), or including participants younger than 45 years old (n=1). A total of 42 articles were eligible for inclusion. The study selection flow diagram is shown in Figure 1.

**Figure 1.** PRISMA flow diagram of reviewed publications and results

Of the 42 studies, 26 (62%) reported some ethno-racial identity data. The 42 eligible studies included a total of 100,748 participants, including the 76365 participants in studies that reported ethno-racial information. White race was reported in 23 trials, Black or African American in 15, Asian in 6, American Indian or Alaska Native in 3, Hawaii Native or Pacific Islander in 1. No studies reported bi-racial identity. In 9 of these trials, White race was explicitly centered and either no other race categories were listed, or all other races were combined into an “Other Race” category. In one instance, only the African American proportion of the sample was reported and all other races including White were captured under “Other Race.” Hispanic ethnicity was reported in 10 trials, including two trials for only Hispanic individuals. In all studies, Hispanic ethnicity appeared to be included as a separate ethno-racial category with no intersection with an identified racial identity. No studies reported enrolling exclusively White, non-Hispanic participants.

Table 1 shows the characteristics of the eligible studies stratified by whether the studies reported ethno-racial information or not. Studies that reported ethno-racial information did not have different sample sizes (p=0.48, CI [-5459.7, 2633.4]), average age (p=0.75, CI [-5.4, 3.9]), or have a different percentage of women (p=0.24, CI [-13.9, 3.5]). Studies that reported ethno-racial information did not employ different intervention types (X^2^ = 8.0, p=0.33), or have significantly different funding (X^2^ = 9.8, p=0.08) but in general were more often funded by the National Institutes of Health (62% vs 19%). Only two studies were explicitly delivered in Spanish, both of which exclusively enrolled individuals who identified as Hispanic.

**Table 1.**
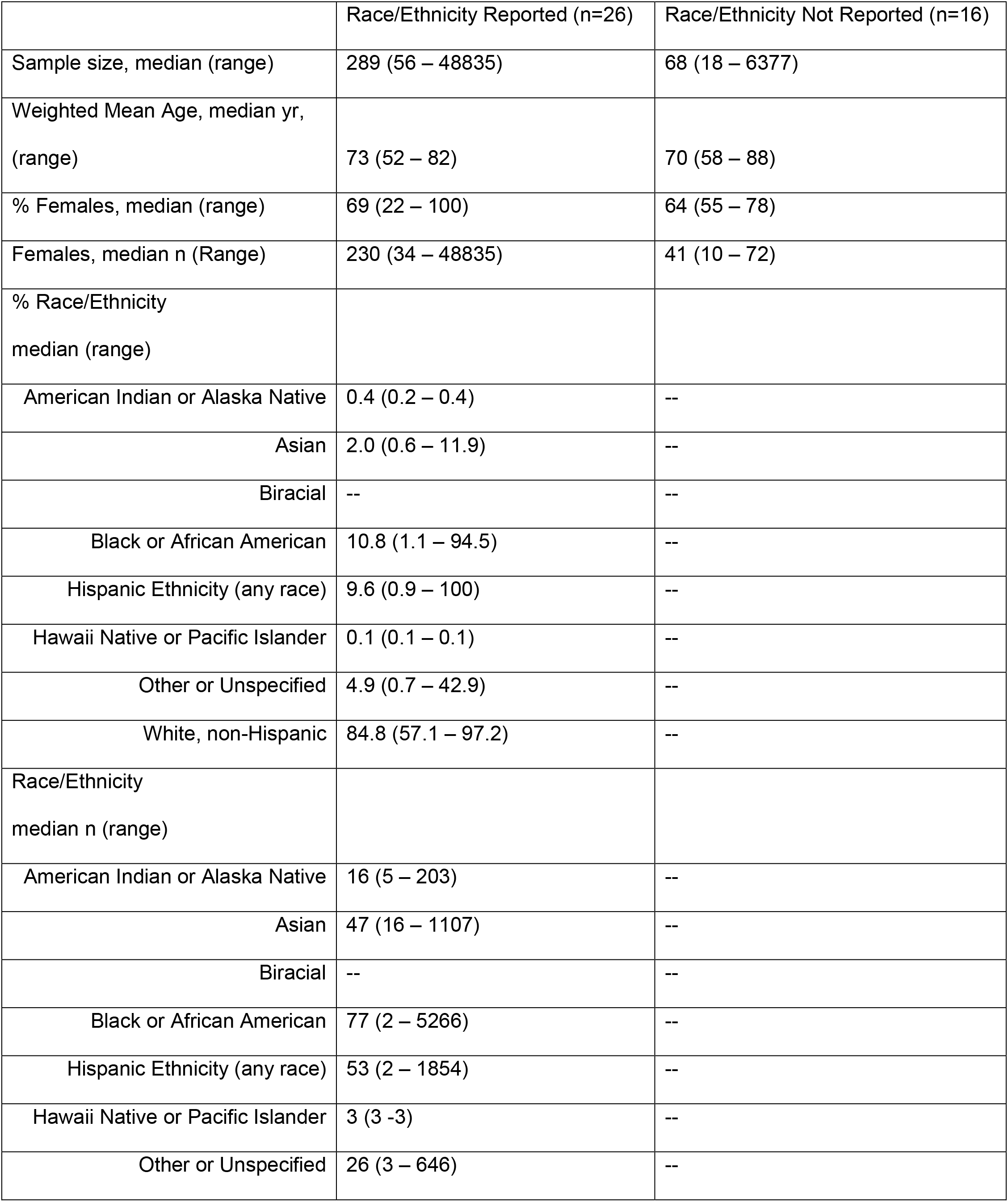
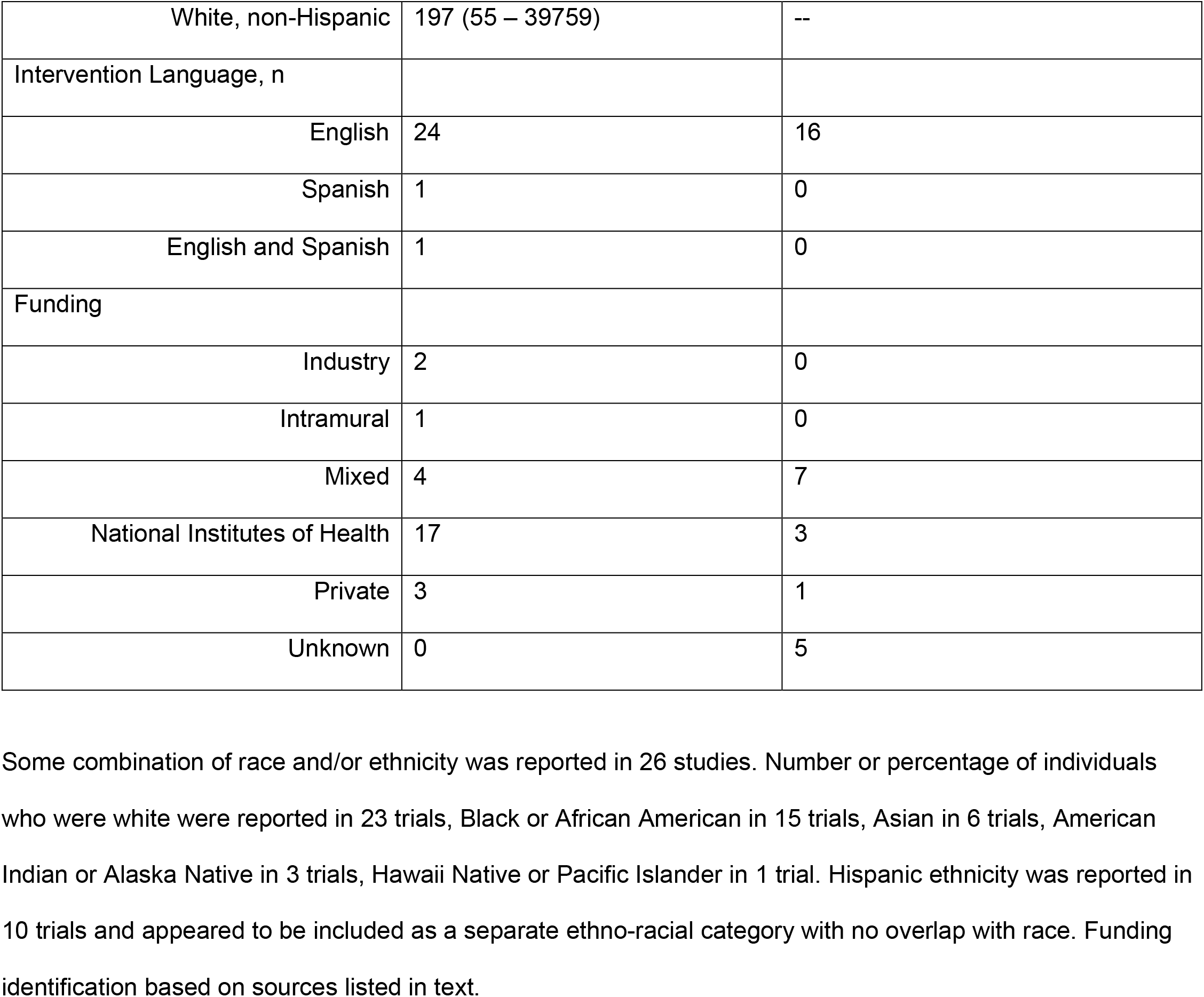
Characteristics of Eligible Studies

The overall and subgroup frequency of each level and the pooled estimated proportions of participants from ethno-racial minority groups are illustrated using forest plotting, Figure 2. The plot displays the estimate and confidence intervals for both overall and subgroup analyses. For studies that reported racial or ethnic identity of participants, the estimated pooled percentage of REM participation was 25.6%. Only the type of intervention demonstrated differences in the proportion of ethno-racial minorities in the sample, (p<0.003). Specifically, diet studies had a lower estimated pooled proportion of REM enrollees. There was insufficient evidence to conclude the proportion of minority enrollees different by funding source or cognitive measure employed.

**Figure 2.** Pooled Proportion of REM in brain healthy prevention trials between 2004 and 2020 using meta-analyses, and subgroup analyses to assess heterogeneity.

## DISCUSSION

In the current study, we conducted a systematic review to characterize the number of REMs enrolled in brain health and prevention trials. We conducted this research because inclusion of REMs in dementia prevention studies is vital to reducing the impact of disparities in dementia risk and critical for addressing imperative gaps in knowledge. Our findings suggest that, between 2004 and 2020, one third of studies failed to report ethno-racial information. Of the studies that did report this information, we found the estimated pooled percentage of REM participation was 25.6%.

This estimate is higher than the representation of REMs in dementia pharmacological treatment RCTs as reported in a 2007 review(18), 10% in NIH and 3.2% in industry-funded RCTs. However, it is important to note that one third of studies in the current review did not report race or ethnicity, which suggests that the numbers of minorities are likely low. Also, 25.6% is lower than the 2019 the U.S. Census Bureau estimate of 30.4% people 45 years and older that identified as a REM, demonstrating that REM remain under-represented in preventative RCTs.(19) Our study findings of the lack of representation of REM in brain health and prevention trials, aligns with current evidence demonstrating the underrepresentation of culturally and linguistical populations in clinical trials.(20-22) Lack of racial/ethnic representation in clinical trials is problematic for generalizability of study findings and equity for REM in obtaining the benefits of participating in clinical trials.(23) It is also troubling given that at a minimum, we know that individuals identifying as African American, Hispanic, and American Indians are at greater risk of developing cognitive change.(24, 25)

Barriers to REM participation in clinical trials are well-documented (26, 27) due to past unethical research practices that have had a significant influence on the community resulting in mistrust of research, medical, and academic institutions as a whole.(28, 29) Additionally, cultural and institutional barriers have impacted REM participation including primary use of passive recruitment strategies (e.g. flyers) that are not culturally tailored to REM communities. Study designs and inclusion criteria for clinical trials also present significant challenges for REM participation (e.g. medical/health eligibility criteria, time/duration of study, transportation requirements to go to study sites).(11, 13, 30)

The prevalence of dementia is high among REM, especially among older African Americans (13.8%) and Hispanics (12.2%) compared to Whites (10.3%).(31) Although evidence consistently indicates that disparities in dementia disproportionately impact REM, especially in terms of incidence, prevalence, diagnosis, and disease burden, the same populations have been historically underrepresented and nearly absent in dementia research,(14, 18, 25, 32) in which less than 4% of ADRD prevention brain health trials are focused on REM communities.(33) However, this review found that an estimated 25.6% enrollment of REM in dementia prevention trials, which indicates there have been improvements made within research to enhance participation and inclusion of REM in clinical trials.

Effective approaches to increase recruitment among REM in research have been made, which emphasize the importance of forming sustainable partnerships with REM communities (e.g. community centers, churches, and trusted community leaders). Centering the community partnerships at all phases of the research in useful to acquiring buy-in from the community as a whole.(34) For example, a church-based HIV intervention used community based participatory research approach to increase HIV testing among African Americans resulting in increased HIV testing (59% vs. 42%, p = 0.008) and church-based testing (54% vs. 15%, p < 0.001) within 12 months.(35) In another in trial, Promotoras de Salud delivered a diabetes prevention program for prediabetic Latina adults in Spanish. Participants reported overall high satisfaction with the culturally tailored program and results indicated significant reduction in weight loss (5.6% of initial body weight) and cardiovascular related risk factors (e.g. diastolic blood pressure, insulin, and LDL cholesterol).(36) These trials demonstrate that community driven trials that are culturally tailored result in feasibility, acceptability, and effectiveness in addressing health disparities among REM. Therefore, it is imperative that dementia prevention trials involve community partners to enhance recruitment and acceptability among REM. In addition, resources through the NIA’s Office of Special Populations are available to support recruitment and retention efforts for REM.(37) Policy-related strategies might also help increase the representation of REMs in dementia prevention studies. The NIA is considering a funding strategy for Practice Based Research Networks, which have been reported to increase access to a more diverse population.(38) Another policy may be ensuring accountability by means of contingencies upon achieving the quotas of REM participation proposed in the research design. The NIA Clinical Research Operations & Management System (CROMS), currently being piloted, may assist track recruitment and retention to make it more equitable.(39) Providing incentives has also been shown to enhance participation in dementia clinical trials. African Americans reported that receiving financial compensation, cognitive and genetic tests results would make them more likely to enroll in dementia focused clinical trials,(28) demonstrating that cultural alignment and the use of incentives can support strides towards achieving representation in dementia prevention trials.

This study incorporated a comprehensive systematic methodological search using both electronic databases and gray literature. This study had a few limitations which merit discussion. First, we applied a U.S. based study limit to our review to focus on U.S. based REMs which limits generalization of findings to REMs outside of U.S. Second, we limited our search to studies reported in English, which has been argued to result in systematic bias.(40) Third, we included only studies that reported change in cognitive scores, therefore excluding those that only reported dementia or MCI incidence as an outcome. Future research should explore those studies, although there are likely few, due to the required long time periods to assess those outcomes. In conclusion, this systematic review highlights the lack of ethno-racial reported among participants in brain health prevention RCT trials. Representation of REM is dementia prevention trials is critical to reducing the disproportionate burden dementia has among these populations. Reporting of ethno-racial within dementia prevention trials is encouraged and use of effective recruitment including collaboration with community partners is suggested to enhance recruitment for future dementia prevention trials.

## Supporting information

Figure 1. PRISMA Flow Diagram

Figure 2. Pooled Proportion of REM in brain healthy prevention

Table 1. Characteristics of Eligible Studies

## Data Availability

All data will be de-identified and available for other investigators at their request, in compliance with the NIH guidelines

## ACKNOWLEDGEMENTS

Not applicable.

