## Supplementary figures and images for "Representation of Racial and Ethnic Minority Populations in Dementia Prevention Trials: A Systematic Review"

### Figure 1. PRISMA Flow Diagram

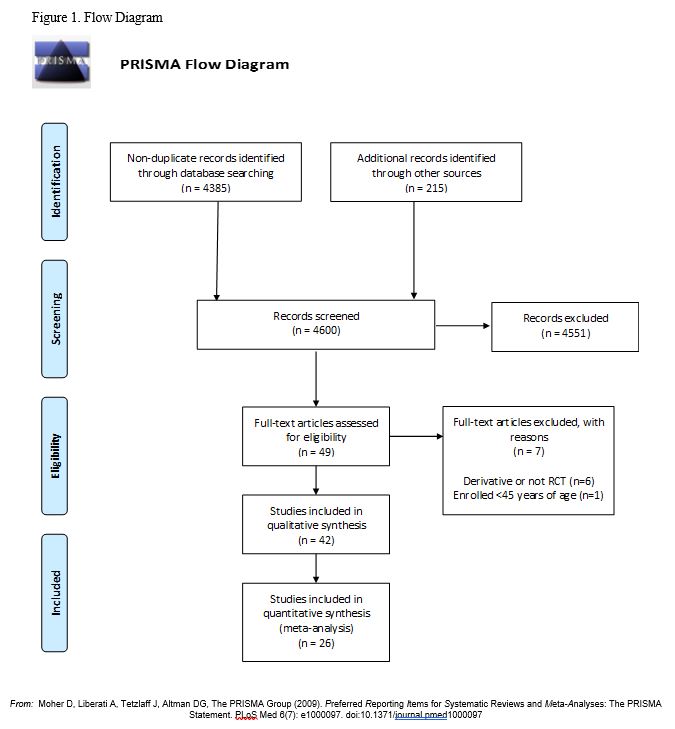

### Figure 2. Pooled Proportion of REM in brain healthy prevention

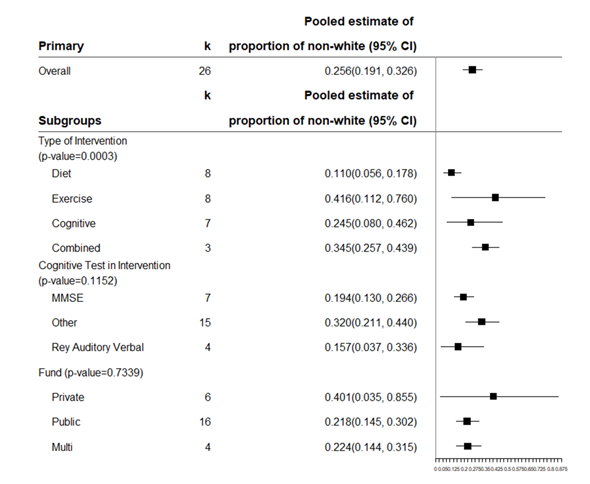
