## Supplementary material for "Representation of Racial and Ethnic Minority Populations in Dementia Prevention Trials: A Systematic Review": Table 1. Characteristics of Eligible Studies

| Table 1 | Race/Ethnicity Reported (n=26) | Race/Ethnicity Not Reported (n=16) |
|---|---|---|
| Sample size, median (range) | 289 (56 – 48835) | 68 (18 – 6377) |
| Weighted Mean Age, median yr, (range) | 73 (52 – 82) | 70 (58 – 88) |
| % Females, median (range) | 69 (22 – 100) | 64 (55 – 78) |
| Females, median n (Range) | 230 (34 – 48835) | 41 (10 – 72) |
| % Race/Ethnicity median (range) | | |
| American Indian or Alaska Native | 0.4 (0.2 – 0.4) | -- |
| Asian | 2.0 (0.6 – 11.9) | -- |
| Biracial | -- | -- |
| Black or African American | 10.8 (1.1 – 94.5) | -- |
| Hispanic Ethnicity (any race) | 9.6 (0.9 – 100) | -- |
| Hawaii Native or Pacific Islander | 0.1 (0.1 – 0.1) | -- |
| Other or Unspecified | 4.9 (0.7 – 42.9) | -- |
| White, non-Hispanic | 84.8 (57.1 – 97.2) | -- |
| Race/Ethnicity median n (range) | | |
| American Indian or Alaska Native | 16 (5 – 203) | -- |
| Asian | 47 (16 – 1107) | -- |
| Biracial | -- | -- |
| Black or African American | 77 (2 – 5266) | -- |
| Hispanic Ethnicity (any race) | 53 (2 – 1854) | -- |
| Hawaii Native or Pacific Islander | 3 (3 -3) | -- |
| Other or Unspecified | 26 (3 – 646) | -- |

| | | |
|---|---|---|
| White, non-Hispanic | 197 (55 – 39759) | -- |
| Intervention Language, n | | |
| English | 24 | 16 |
| Spanish | 1 | 0 |
| English and Spanish | 1 | 0 |
| Funding | | |
| Industry | 2 | 0 |
| Intramural | 1 | 0 |
| Mixed | 4 | 7 |
| National Institutes of Health | 17 | 3 |
| Private | 3 | 1 |
| Unknown | 0 | 5 |

Some combination of race and/or ethnicity was reported in 26 studies. Number or percentage of individuals who were white were reported in 23 trials, Black or African American in 15 trials, Asian in 6 trials, American Indian or Alaska Native in 3 trials, Hawaii Native or Pacific Islander in 1 trial. Hispanic ethnicity was reported in 10 trials and appeared to be included as a separate ethno-racial category with no overlap with race. Funding identification based on sources listed in text.
